# Risk interactions of coronavirus infection across age groups after the peak of COVID-19 epidemic

**DOI:** 10.1101/2020.05.17.20105049

**Authors:** Xinhua Yu

## Abstract

**Background:** the COVID-19 pandemic has incurred significant disease burden worldwide, particularly on elderly population. This study aims to explore how risks of infection interact across age groups using data from South Korea.

**Methods:** Daily new COVID-19 cases from March 10 to April 30, 2020 were scraped from online open sources. A multivariate vector autoregressive model for time series count data was used to examine the risk interactions across age groups. Case counts from previous days were included as predictors to dynamically examine the change of risk patterns.

**Results:** In South Korea, the risk of coronavirus infection among elderly people was significantly affected by other age groups. An increase of virus infection among people aged 20-39 was associated with a double risk of infection among elderly people. Meanwhile, an increase in virus infection among elderly people was also significantly associated with risks of infection among other age groups. The risks of infection among younger people were relatively unaffected by that of other age groups.

**Conclusions:** Protecting elderly people from coronavirus infection could not only reduce the risk of infection among themselves but also ameliorate the risks of virus infection among other age groups. Such interventions should be effective and for long term.

## 1. Introduction

The coronavirus disease 2019 (COVID-19) is caused by the infection of a novel Severe Acute Respiratory Syndrome associated coronavirus (SARS-CoV-2) [1]. Since December 2019, over 11 million people have been infected with SARS-CoV-2 and over 528,000 people died of coronavirus infection (https://coronavirus.jhu.edu/map.html, accessed on July 4, 2020). Of them, elderly people and people with underlying chronic conditions suffered the heaviest disease burden [2–4]. For example, about 80% of deaths were people aged 65 or above (https://www.cdc.gov/coronavirus/2019-ncov/cases-updates/cases-in-us.html), and 43.4% of hospitalizations aged 65 or above [5]. In the state of Florida, US, people aged 65 or above accounted for 54% of hospitalizations, and the mortality rate was 14% if infected with virus [6].

The reasons for the disproportional burden among elderly people were unclear [7]. Elderly people generally have weaker immune system than younger people due to aging, and they are also more likely to have multiple chronic conditions [8,9]. Thus, elderly people may have severe symptoms if infected with coronavirus [10,11]. On the other hand, elderly people may have exposed myriads of infections over their lifetime which may provide immunity against new virus infection. Although cross-reaction of antibodies between SARS-CoV and SARS-CoV-2 was observed, cross-neutralization was rare [12]. Thus, it was unlikely elderly people might have any effective immunity against SARS-CoV-2.

Meanwhile, the COVID-19 pandemic is waning down in some countries such as South Korea since March 10, 2020 (see Figure 1) [13,14], and society is gradually returning to normalcy [15]. A potential rebound of new cases has been warned by many public health experts [16]. This is reflected in an epidemic curve with a long tail and occasional spikes, which is demonstrated in the epidemic process in South Korea (https://www.kcdc.info/covid-19/) [17,18]. In addition, if the seasonality, immunity and cross-immunity of SARS-CoV-2 behave like previous coronaviruses, a recent study predicted a long lasting and multi-wave epidemic was possible in the US [19]. Therefore, it is imperative to examine risk patterns of coronavirus infection among elderly people after the peak of epidemic.

**Figure 1.**
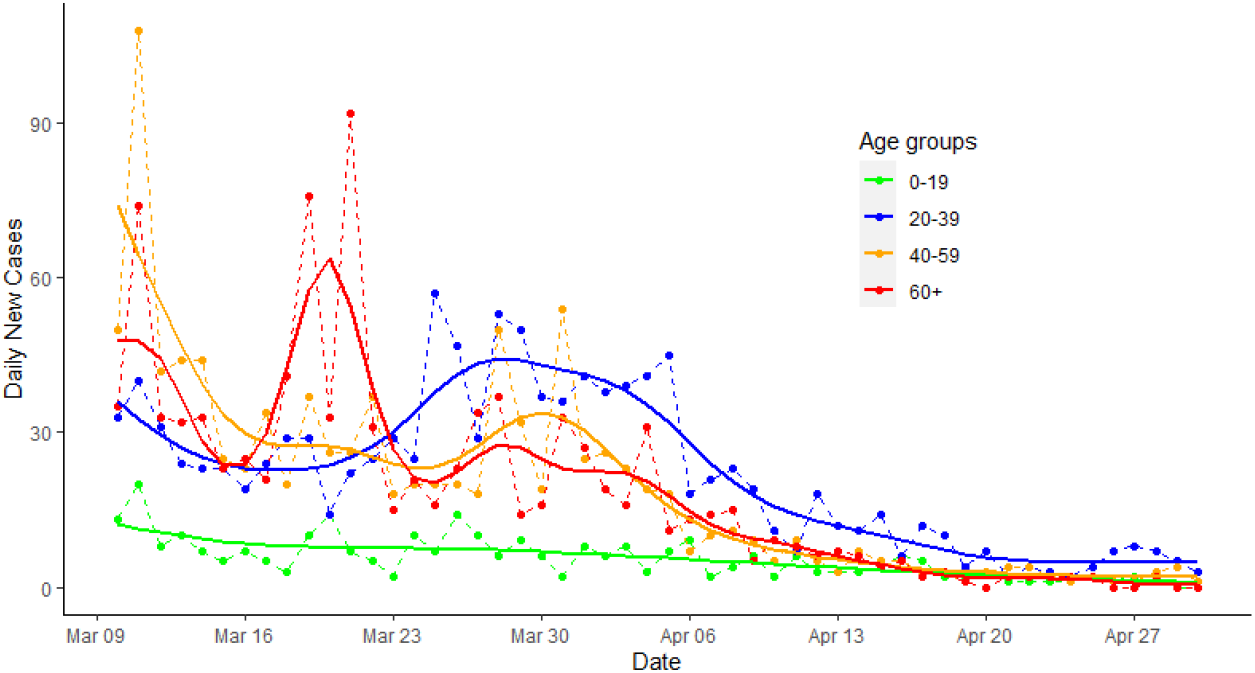
Epidemic curve of COVID-19 in South Korea from March 10 to April 30, 2020, by age groups

Unfortunately, due to lack of testing kits and heterogeneous diagnosis criteria, epidemiological data on COVID-19 among different countries (and even within a country) were often noncomparable [20]. One notable exception is South Korea where extensive contact tracing and mass testing not only curtailed the epidemic but also generated high quality data. In South Korea, both asymptomatic and symptomatic cases were identified promptly [17,18]. Thus, a complete picture of the epidemic process was possible to depict.

In this study, we will examine how risks of coronavirus infection interact across age groups using time series analysis. Using the high quality data from South Korea, we will focus on the post-peak period of the epidemic process to evaluate the risk of infection among elderly people during the period of society re-opening.

## 2. Materials and Methods

Daily new COVID-19 case counts from South Korea were obtained from the website (https://www.kaggle.com/kimjihoo/coronavirusdataset) which were scraped from the Korea Center for Disease Control website. The first COVID-19 case in South Korea appeared on Jan 20, 2020, and the major epidemic started on Feb 19, 2020. Since the peak of first epidemic wave in South Korea ended around March 10 [21] (also Figure 1), we limited the time series of new cases between March 10 to April 30, 2020 for South Korea. All daily cases were stratified by age groups (0-19, 20-39, 40-59, and 60 or above). Those aged 60 or above were referred as elderly people.

The observed epidemic curves by age groups from March 10 to April 30, 2020 were plotted, and the predicted daily cases were obtained with a generalized additive model (GAM) [22] assuming daily new cases follow Poisson distributions. The smoothness of predicted values was achieved with thin plate regression splines with 16 knots using R *mgcv* package (see Appendix A).

We developed a vector autoregressive (VAR) model to examine the associations of the infection risks across age groups simultaneously [23]. Specifically, we assumed daily new case counts (y_j,t_) followed a generalized Poisson distribution to account for over-dispersion of case counts (i.e., observed variance is larger than expected variance) [24]. The model also included case counts from previous days (lags) across age groups as predictors to form a dynamic model (see Appendix A for details). Therefore, the current risk of infection in each age group was predicted not only by previous case counts in its own group but also by previous counts from other age groups.

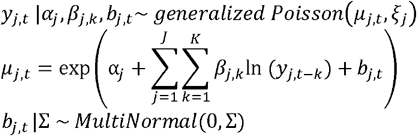

Where *j* = *1*,…,*J* represented age groups, *t*=*1*,…,*T* represented days, and *k* = *1*,…,*K* represented the number of time lags. Because the typical incubation period of COVID-19 is five days [25], we reported results from five-lag models. Three-lag and seven lag models were also explored, and results from all models were consistent (see online codes and results). The scale parameter *ξ* in the generalized Poisson distribution controls the magnitude of dispersion, that is, *ξ* = 0 corresponding to a standard Poisson (mean = variance), *ξ* < 0 suggesting under-dispersion (mean > variance), and 0 < *ξ* < 1 indicating overdispersion (mean < variance). The *b*_*j,t*_ could be viewed as a random effect to account for the correlation of daily counts between age groups. The *b*_*j,t*_ was assumed a multivariate normal distribution.

The above model framework was similar to the common log-linear relative risk models in epidemiological studies which assume multiplicative associations between predictors and outcomes [26]. The coefficients βs could be interpreted as natural logarithms of risk ratios per one unit change of natural logarithms of case counts.

We fit the above models with Bayesian software stan through Rstan interface (http://mc-stan.org) [27]. To keep the model simple, we assumed weakly informative priors of student t distributions for all αs and βs, and an LKJ prior with modal density around diagonals for correlations between case series (see Appendix A). Hamiltonian Monte Carlo was used to obtain posterior distributions of parameters. Diagnostic plots showed all chains mixed satisfactorily and were converged. In addition, negative binomial models were also fitted, and results were similar to those reported here except for wider confidence intervals (Appendix B, Tables 1). Noticed that the dispersion factors estimated from the generalized Poisson models were 2.66 (1.58 - 5.13), 1.44 (0.96 - 2.46), 2.21 (1.37 - 3.82), and 0.90 (0.59 - 1.50) for those aged 60 or above, 40-59, 20-39, and 0-19, respectively. These estimates were of moderate magnitude and two of them (age group 40-59 and 0-19) were not statistically different from 1. This also suggested that negative binomial models might overestimate the dispersion factors which led to wider confidence intervals. The data, replicable codes and other results were available online (www.github.com/xinhuayu/riskinteractions/).

**Table 1.** Risk interactions in coronavirus infection across age groups during the COVID-19 pandemic, South Korea, March 10 to April 30, 2020

| Model |  | Lags of predictors |  |  |  |  |
| --- | --- | --- | --- | --- | --- | --- |
| Outcomes | Predictors | Lag 1 | Lag 2 | Lag 3 | Lag 4 | Lag 5 |
| Aged 60 or above |  |  |  |  |  |  |
|  | 60 or above | 2.09 (1.28 - 3.17) * | 1.21 (0.78 - 1.80) | 0.90 (0.60 - 1.39) | 1.03 (0.60 - 1.67) | 0.96 (0.63 - 1.53) |
|  | 40 - 59 | 0.95 (0.49 - 1.84) | 1.81 (0.98 - 3.29) | 0.59 (0.30 - 1.18) | 1.37 (0.85 - 2.23) | 1.03 (0.61 - 1.70) |
|  | 20 - 39 | 0.89 (0.53 - 1.46) | 2.02 (1.12 - 3.47) * | 1.28 (0.72 - 2.31) | 0.41 (0.22 - 0.78) # | 1.11 (0.58 - 2.22) |
|  | 0 - 19 | 0.90 (0.61 - 1.43) | 1.36 (0.95 - 1.95) | 0.82 (0.57 - 1.19) | 0.61 (0.44 - 0.85) | 1.64 (1.03 - 2.58) * |
| Aged 40 - 59 |  |  |  |  |  |  |
|  | 60 or above | 1.66 (1.19 - 2.29) * | 0.79 (0.55 - 1.12) | 0.89 (0.64 - 1.23) | 0.98 (0.70 - 1.40) | 1.02 (0.71 - 1.40) |
|  | 40 - 59 | 1.01 (0.62 - 1.63) | 1.76 (1.16 - 2.66) * | 1.23 (0.76 - 1.96) | 0.89 (0.62 - 1.29) | 0.88 (0.62 - 1.28) |
|  | 20 - 39 | 1.13 (0.75 - 1.73) | 1.12 (0.76 - 1.67) | 1.59 (1.03 - 2.50) * | 0.60 (0.37 - 0.95) | 0.88 (0.53 - 1.48) |
|  | 0 - 19 | 1.04 (0.79 - 1.37) | 1.28 (0.97 - 1.68) | 1.17 (0.88 - 1.57) | 1.14 (0.88 - 1.49) | 1.17 (0.84 - 1.62) |
| Aged 20 - 39 |  |  |  |  |  |  |
|  | 60 or above | 0.95 (0.69 - 1.29) | 0.89 (0.65 - 1.23) | 1.01 (0.72 - 1.39) | 1.54 (1.11 - 2.12) * | 0.97 (0.69 - 1.31) |
|  | 40 - 59 | 1.17 (0.73 - 1.88) | 1.18 (0.78 - 1.78) | 0.98 (0.60 - 1.52) | 0.90 (0.64 - 1.26) | 1.04 (0.75 - 1.47) |
|  | 20 - 39 | 1.56 (1.05 - 2.36) * | 1.02 (0.68 - 1.55) | 1.45 (0.96 - 2.24) | 1.06 (0.68 - 1.64) | 0.85 (0.55 - 1.32) |
|  | 0 - 19 | 1.04 (0.79 - 1.37) | 0.96 (0.75 - 1.27) | 0.88 (0.66 - 1.20) | 0.97 (0.75 - 1.26) | 0.74 (0.54 - 1.00) |
| Aged 0 - 19 |  |  |  |  |  |  |
|  | 60 or above | 1.78 (1.23 - 2.61) * | 1.34 (0.86 - 2.06) | 0.99 (0.67 - 1.51) | 0.82 (0.52 - 1.25) | 1.55 (1.02 - 2.30) * |
|  | 40 - 59 | 0.76 (0.41 - 1.38) | 0.71 (0.42 - 1.18) | 0.85 (0.47 - 1.56) | 0.91 (0.55 - 1.57) | 0.97 (0.62 - 1.54) |
|  | 20 - 39 | 1.51 (0.92 - 2.55) | 1.50 (0.85 - 2.57) | 1.04 (0.61 - 1.80) | 0.78 (0.44 - 1.38) | 0.91 (0.51 - 1.62) |
|  | 0 - 19 | 0.93 (0.64 - 1.36) | 0.85 (0.60 - 1.21) | 0.85 (0.60 - 1.27) | 0.92 (0.62 - 1.38) | 1.51 (0.99 - 2.31) |

Note: \* and # for p < 0.05

### Ethics statement

This study was based on publicly available data. There was no direct involvement of human subjects. Therefore, it was exempted from the approval of Institutional Review Board. No informed consent was needed. All authors declared no conflict of interest in conducting this study.

## 3. Results

In South Korea, there were 3,383 COVID-19 cases between March 10 and April 30, 2020. Of them, 283 cases aged 0-19 (8.4%), 1,141 aged 20-39 (50.0%), 987 aged 40-59 (29.2%), and 972 aged 60 or above (28.7%).

Figure 1 presented the epidemic curves with fitted values by age groups for South Korea. After March 10, there was a small pike among those aged 60 or above around March 20,2020, and a small rebound among those aged 20-39 (e.g., around March 30 to April 5, 2020), followed by those aged 40-59 and aged 60 or older.

Table 1 described associations of risks of infection across age groups in South Korea. In addition to tracking effect from the first lag day (yesterday) among elderly people, the current risk of infection among elderly people was associated with double risk by one unit increase of infection risk among those aged 20-39 in the second lag day. Additionally, an increase of infection in the youngest population during the fifth lag days was also associated with an increased risk of infection among elderly people by 64%.

More importantly, an increase of virus infection among elderly people was associated with increases in risks of infection among all other age groups, but with longer delays in younger populations. Furthermore, the risk of infection among people aged 40-59 was affected by both old and young people, but to a less extent. Risks of infection among people aged 20-39 or 0-19 were less likely affected by other age groups.

## 4. Discussion

This was the first study to quantify risk interactions of SARS-CoV-2 infection across age groups based on vector autoregressive models using epidemic data of high quality from South Korea. We found that in South Korea, the risk of infection among elderly people was significantly affected by other age groups. An increase in virus infection among elderly people was also significantly associated with increased risks of infection among other age groups. Risks of infections among younger people were relatively unaffected by that of other age groups.

Our results were consistent with the current COVID-19 epidemic process, in which risk of infections among elderly people might be affected by other age groups [28]. Although virus transmission might differ among age groups [7,29], the risk interactions were likely due to personal interactions between people of different age groups. Respiratory infectious diseases often spread through personal contacts [30]. Previous studies showed that contacts were more frequent in young age groups than older age groups, and interactions across age groups were less frequent than within each age group [31]. During the emerging pandemic like COVID-19, stringent control measures such as lock-down, strict social distancing and stay-at-home rules, were often implemented promptly, leading to a significantly abrupt change of contact patterns within and between age groups. Modern techniques such as contract tracing app, infection risk ID, and instant notification of cases, also allowed us to efficiently isolate cases and quarantine high risk people. The observed risk interactions between age groups in South Korea might be largely due to the change of contact patterns during the epidemic period. As shown in our study, there were 2-5 lagging days in the risk interactions across age groups, especial between old and young people. On the other hand, the infection among elderly people may still be affected by and also affect the risks of infection among other age groups. Passive community interactions such as grocery shopping might play an important role in sustaining the epidemic.

Our results highlighted the importance of implementing and enforcing effective interventions in the whole society [32–34], and the highest priority of protecting elderly people [29]. Furthermore, we showed that an increase of coronavirus infection among elderly people was associated with increased risks of infection among other age groups, suggesting protecting elderly people and reducing the risk of infection among elderly people had spillover effect in the whole society. This was consistent with our previous simulation study in which reducing contacts among elderly could reduce the virus infection and hospitalizations in the whole society [35]

There were some limitations in this study. The most important one was that we relied on reported cases. The data from South Korea were more likely complete due to extensive contact tracing and mass testing. Furthermore, the case reporting date (or virus infection detection/lab confirmation date) was different from the virus infection date, and the average incubation period for SARS-CoV-2 was about 5 days [25]. The laudable efforts of extensive contact tracing and mass testing implemented by the South Korea government at the beginning of COVID-19 epidemic significantly reduced the reporting delays, and likely identified many cases before symptom onsets [17]. Therefore, the interval between virus infection and case reporting might be small. In addition, there were other factors such as gender, socio-economic status and neighborhood environment might also affect the risk of infection.

Moreover, although we interpreted the results with action terms, they had no explicit causative meanings. For example, younger people tended to have milder or no symptoms (i.e., subclinical cases) if infected with virus [36–38]. Thus, it was possible that an increased number of detected cases among young people implied the existence of an increase in subclinical cases in the community who might unknowingly infect other people, including elderly people. Subclinical cases could only be identified through extensive contact tracing and mass testing. Without this information, it is impossible to examine the route of infections in the community.

Our study has several strengths. Firstly, data from South Korea were more likely complete which would provide information about underlying epidemic mechanisms. Although different social norms and health care systems might explain some differences in risk patterns between South Korea and other regions, results from South Korea provided a baseline picture of risk interactions among age groups under a well-controlled, ideal epidemic process. Different patterns might be due to differences in population structures, magnitudes of control measures and contact patterns in the society, while similar patterns in risk interactions between regions allowed us to infer the possible paths of infections.

Secondly, we proposed a novel multivariate autoregressive model for time series of counts to examine the risk of virus infection across age groups simultaneously. A flexible generalized Poisson model fitted with Bayesian methods was used to account for overdispersion of count data [24]. Unlike many other studies that used mechanistic epidemic models which was useful to describe the epidemic process [39], our statistical models extended traditional relative risk models to time series of count data. It should be note that this type of model likely overfit the data and collinearity among lag variables also exist. Thus, having *a priori* hypotheses and choosing biologically relevant lags are critical in building correct models and interpreting the results. Our lag models were based on observed incubation period of COVID-19 and for testing pre-specific hypotheses. The principle of our methods was similar to that of Institute for Health Metrics and Evaluation (IHME) [40] and University of Texas-Austin models [41], all of which relied on time series analysis of count data. However, we did not attempt to predict future cases. Rather, we focused on disentangling risk interactions of infection across age groups, which was more important and relevant in disease preventions.

Finally, during the process of re-opening the economy and society, the number of new cases may rebound, multiple small waves or a second big wave of epidemic are possible. A contentious issue was whether and how to protect high risk populations such as elderly people during the return of epidemic. Therefore, we limited our study period to the post-peak of epidemic to answer this imminent question. Our study strongly supported that high risk populations such as elderly people should still take serious precautions during the post-epidemic period.

## 5. Conclusions

In summary, protecting elderly people from coronavirus infection might not only be associated with a reduced risk of infection among themselves but also related to lower risks of virus infection among other age groups. Therefore, elderly people should keep on practicing social distancing and maintaining effective personal protections until the pandemic is completely over.

## 6. Patents

N/A

## Data Availability

GitHub address later

## Supplementary Materials

N/A

## Author Contributions

Dr. Xinhua Yu has full access to research data and conducted data analysis and report writing.

## Abbreviations

The following abbreviations are used in this manuscript:

SARS-CoV-2: severe acute respiratory syndrome coronavirus 2
COVID-19: coronavirus infectious disease 2019
GAM: generalized additive model
VAR: vector autoregressive regression
NB: negative binomial model
REML: restricted maximum likelihood
PMF: probability mass function

## Appendix A

### I. Obtained Smoothed predicted daily cases with Generalized Additive Model (GAM)

Assuming daily new cases follow a Poisson distribution or negative binomial (NB) distribution (see below), the GAM is a linear regression with smoothed time term. For simplicity, a separate GAM regression was fitted for each age group:

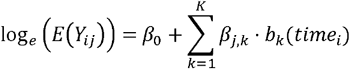

where Y_*ij*_ represents the observed case counts of day *i* and group *j*, and E(Y_ij_) is expected (predicted) value. The variable timei represents day (1,…,I), *b*_*k*_() represents a basis function for the kth term to smooth temporal trend, and β_*j,k*_*s* are regression coefficients for smooth term *k* and group *j*. Restricted maximum likelihood (REML) approach was used in parameter estimation. R mgcv package was used [22], and smooth terms were fitted using thin plate regression spline with 16 knots.

### II. Model setups and comparisons

The standard Poisson distribution describes the distribution of y events occurring at a constant rate of λ. The Probability mass function (PMF) is:

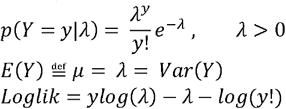

In standard Poisson distribution, expected variance equals mean. If observed variance is larger than expected variance (i.e., the mean), then overdispersion exists. This often occurs when outcomes are correlated, such as daily new case counts during a disease outbreak.

The generalized Poisson distribution introduces an additional scale parameter ξ [42] as quoted in Hilbe JM. 2014 [26]. The PMF is:

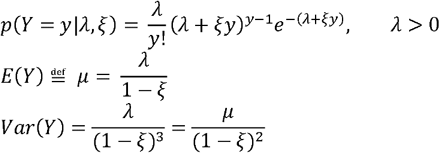

Thus, 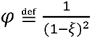 is the dispersion factor indicating how variance changes with the mean.

Therefore, if ξ = 0, then ϕ = 1, corresponds to a standard Poisson (mean = variance); 0 < ξ < 1, then ϕ > 1, models overdispersion (mean < variance); and ξ < 0, then ϕ < 1, models under-dispersion (mean variance).

Reparametrize the PMF of generalized Poisson distribution with μ and ξ[24]:

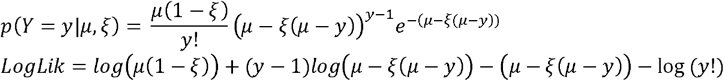

On the other hand, the negative binomial distribution describes the distribution of the number of successes given a predefined r number of failures during a sequence of independent Bernoulli trials with a success probability p:

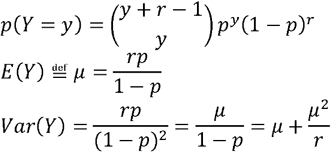

Thus, the overdispersion of Y is controlled by the shape parameter r.

Reparametrize the PMF with μ and r,

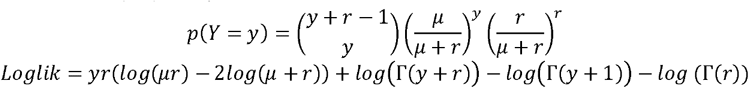

Where gamma function Γ(x+1) = x! (x factorial) for an integer x, and the parameter r can be any positive real value.

Note that negative binomial distribution can be viewed as a Gamma-Poisson mixture distribution in which Y ∼ Poisson(λ) and λ ∼ Gamma(*r*, λ/*r*). That is, the negative binomial distribution (r, p) is the posterior distribution of Poisson(λ) with Gamma(*r*, λ/*r*) as the conjugate prior of λ, where 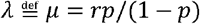 Rewriting Gamma(*r*, λ/*r*) as Gamma(*r,p*/(1− *p*) and using Γ and using Γ functions to represent factorials:

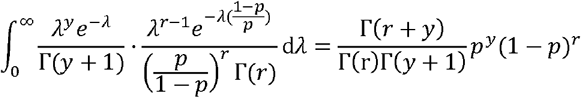

Under this framework, negative binomial distribution is appealing as a natural extension of Poisson distribution to allow for overdispersion that is controlled by the shape parameter *r*.

However, although negative binomial distribution is often used to model new case counts during disease outbreaks, it models only overdispersion and assumes a quadratic relationship between variance and mean, while the generalized Poisson model is more flexible and assumes a simpler first order association between variance and mean. Therefore, we chose to report results from generalized Poisson models. Results from negative binomial models were included in the appendix. In addition, it is also of note that there are extensions of negative binomial models in which the association between mean and variance can be estimated from data, leading to a more flexible model and also permitting the exploration of determinants of overdispersion [26].

In this study, we proposed the following hierarchical vector autoregressive model (VAR) for count data:

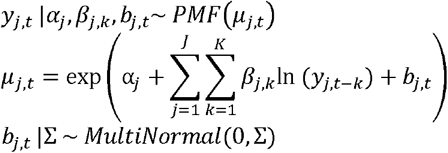

Where *j* = *1*,…,*J* represents age groups, *t*=*1*,…,*T* represents days, and *k* = *1*,…,*K* represents the number of lags. The PMF of Y can be either standard Poisson (λ), generalized Poisson (μ, ξ), or negative binomial (μ, *r*) distribution.

The above VAR model included new case counts from previous days (lags) across age groups as predictors[23], thus examining associations of the infection risks across age groups simultaneously. That is, the current risk of infection in each age group was predicted not only by previous case counts in its own group but also by previous counts from other age groups.

The correlation of daily counts between age groups was modeled through *b_j,t_* that can be viewed as a random effect. The *b_j,t_* was assumed a multivariate normal distribution.

The exponential link between dynamic predictors and μ is equivalent to common relative risk models in epidemiological studies, i.e., log-linear models for count data. Under this multiplicative scale framework, the interpretation of βs are relative risks given one unit increase of predictors.

During the model fitting, we assumed some weakly informative priors for all parameters:

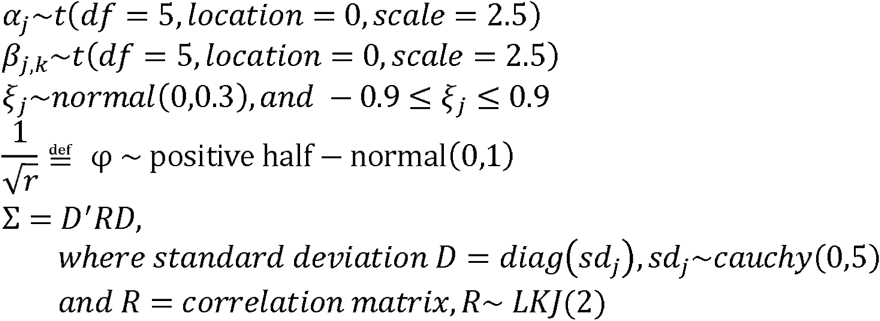

The LKJ prior is a special prior most suitable for correlations. The LKJ(2) assumes a modal density surrounding diagonals.

The models were fit with Bayesian software stan through Rstan interface [27]. A customized stan function was constructed for fitting generalized Poisson model. We employed Hamiltonian Monte Carlo with 5 Markov chains, each with 50,000 iterations plus 2000 warmups, to obtain posterior distributions of parameters. Diagnostic plots through shinestan package showed all chains mixed well and were converged. The replicable data and codes, including models with daily case counts as standard Poisson, generalized Poisson or negative binomial distributions, were available online (www.github.com/xinhuayu/riskinteractions/).

## Appendix B: Additional tables

**Table 1:**
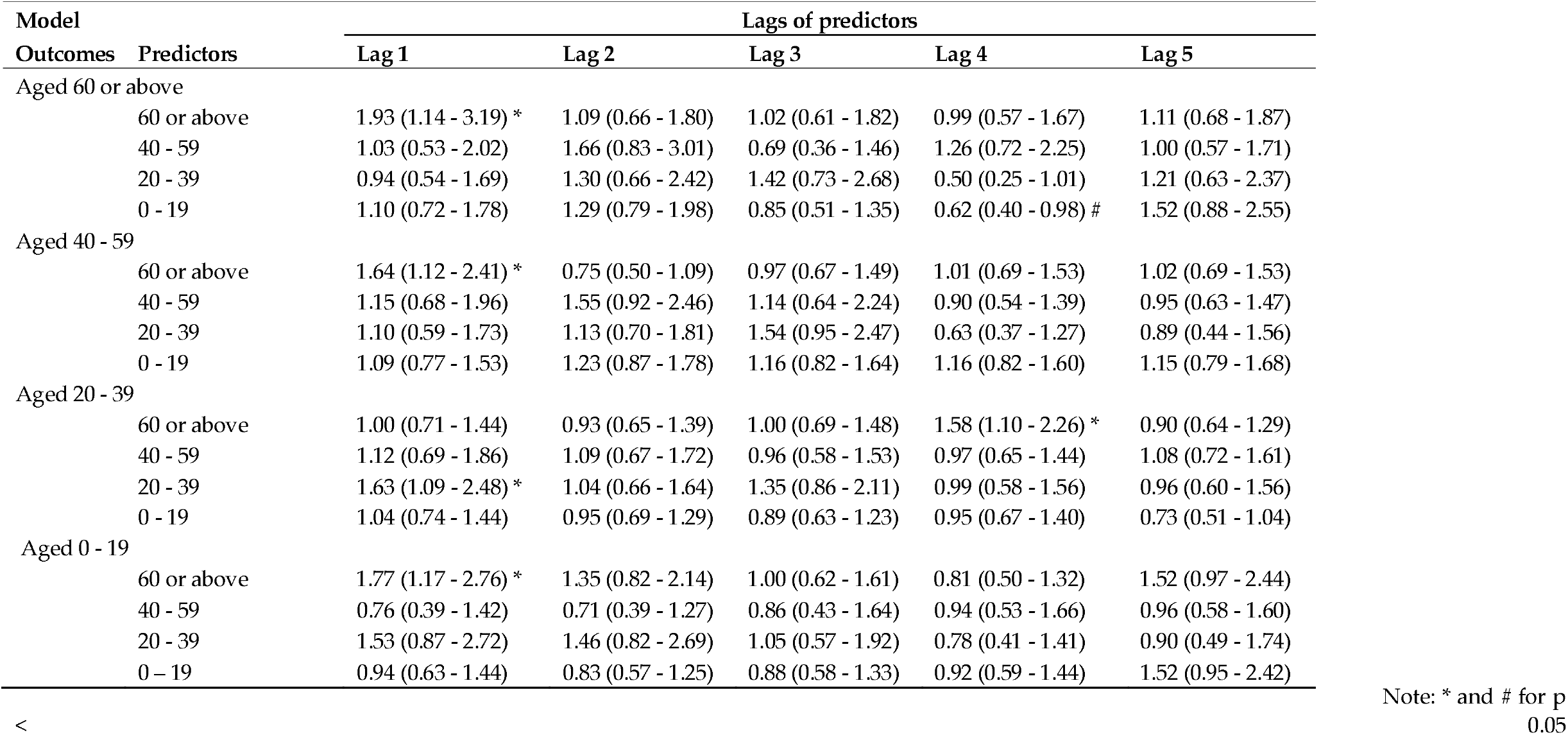
Risk interactions in coronavirus infection across age groups based on negative binomial models, COVID-19, South Korea

